# Research on female breast and gynaecologic cancers in the Democratic Republic of Congo: a scoping review

**DOI:** 10.1101/2024.11.15.24317400

**Authors:** Anifa Kalay, Eric Madiata, Bienvenu Lebwaze, Médard Donzo, John Botomwito, Delly Tshiabo, Mireille L. Ntambwe

## Abstract

**Background:** In the last decades, the incidence of female breast and gynecologic cancers has been increasing in sub-Saharan African countries including in the Democratic Republic of the Congo (DRC). Although DRC has taken some steps in addressing cancer issues, cancer control is not yet efficiently organized. The DRC national strategy for the fight against cervical and female breast cancers stresses that research conducted on female breast and cervical cancers in DRC is not leveraged to support the Ministry of Health. There is a clear and urgent need to identify the type and extend of research conducted in DRC about female breast cancer and cervical cancers to inform the operational cancer research agenda in DRC. Thus, the purpose of this scoping review is to describe current research and identify research gaps related to research on female breast and gynaecologic cancers in DRC

**Methods:** A scoping review was conducted through search of several electronic databases. We included peer-reviewed articles and grey literature resources reporting primary or secondary studies about female breast cancer or gynaecologic cancers in DRC.

**Results:** A total of 448 articles were retrieved. After screening, 32 articles were retained for review. Twenty-one articles pertained to cervical cancer, 12 to female breast cancer and 2 to vulva cancer. Five articles focused on several cancer types. There was no article retrieved for the other gynaecologic cancers (uterine, Fallopian tube and ovarian cancers). Most of the studies (60%) used a cross-sectional design. The studies were conducted in six provinces.

**Conclusion:** This scoping review has demonstrated significant gaps in female breast and gynaecologic cancer research in DRC. The review’s findings support the need for further research in all areas of the continuum of cancer care, the establishment of a clear and adapted research agenda, advocacy and providers’ capacity development.

## Background

In the last decades, the incidence of female breast and most gynaecologic cancers has been increasing worldwide including in sub-Saharan African countries (1,2). In 2020, worldwide female breast cancer, cervical cancer, uterine cancer and ovarian cancer incidence rates were respectively 47.8, 13.3, 8.7, 6.6, per 100 000 persons (3) whereas in 2008 this rates were 39.0, 15.2, 8.2 and 6.3(4).

Gynaecologic cancers develop from women’s reproductive organs including uterine cervix, ovaries, endometrium (uterus), vagina, and vulva (5). Approximately 30% of cervical cancer-related deaths occur in sub-Saharan Africa (6). A study reported that sub-Saharan Africa had the highest female breast cancer -related age-standardized mortality rate (7). A similar conclusion was observed about ovarian cancer (8).

These cancers have a substantial economic impact on women, families, health systems and society because of the cost associated with cancers management and the loss of productivity due the disease (9).

However, there are known prevention practices, screening tests, detection methods, treatment and palliative care approaches that have shown efficiency in reducing morbidity and mortality rates associated with cancer. This has mainly been observed in high income countries. This package of interventions is part of cancer control. Cancer control aims to reduce the incidence, morbidity, and mortality of cancer and to improve the quality of life of cancer patients in a defined population, through the systematic implementation of evidence-based interventions for prevention, early detection, diagnosis, treatment, and palliative care (10).

Nevertheless, cancer control is not efficiently organized in several sub-Saharan African countries including in the Democratic Republic of the Congo (DRC).

In 2020, cervical cancer had the highest estimated cancer-related age-standardized incidence and mortality rates in Congolese women respectively 31.9/100 000 and 23.7 /100 000 (10). Female breast cancer had the second highest rates in DRC, ovarian cancer had the fourth and uterine cancer had the tenth (11).

DRC has taken some steps in addressing cancer control issues. In 2015, the DRC national strategy for the fight against cervical and female breast cancers was developed (12). This strategy highlights screening and diagnosis methods as well as management of cervical and female breast cancer relevant to DRC health system; and identifies five key areas of intervention: communication for behavior change, service delivery, capacity building of human resources, strengthening the management of health information and operational research. The strategy also stresses that research conducted on female breast and cervical cancers in DRC is not leveraged to support the Ministry of Health.

Operational research is also crucial to the new DRC National Center for the Fight against Cancer. The Center, established in November 2020, is responsible for the implementation of the strategy. Thus, operational research will generate data that will inform not only the implementation of the strategy but also monitoring and evaluation activities that are both crucial for cancer control and necessary to the ministry of health and the Center.

There is a clear and urgent need to identify the type and extend of research conducted in DRC about female breast cancer and cervical cancers to inform the operational cancer research agenda in DRC. In addition, other gynecologic cancers cannot be overlooked as they are now a public health issue in Sub-Saharan Africa and hence in DRC.

On the grounds of this, the purpose of this scoping review is to describe current research and identify research gaps related to research on female breast and gynaecologic cancers in DRC.

The following research questions were considered:

1. What is the available research on female breast cancer and gynaecologic cancer in DRC?
2. What are the gaps in female breast cancer and gynaecologic cancer research in DRC?

## Methods

The Joanna Briggs Institute (JBI) approach was used as a framework to guide this scoping review(13). This approach includes five steps: 1) research question definition 2) relevant studies identification 3) study selection 4) data charting 5) results collation, summarization and reporting. The PRISMA extension for scoping reviews (PRISMA-ScR) checklist was used to inform the reporting structure in this review (14).

### Eligibility criteria

Peer-reviewed articles and grey literature resources were included if they reported primary or secondary studies about female breast cancer or gynaecologic cancers in DRC. Only studies published between January 2001 and April 2021 and written in either English or French were included. The most recent search was conducted in June 2021.Quantitative and qualitative studies were considered.

### Information sources

The following databases were consulted to retrieve relevant literature: Medline Ovid, CINHAL, Scopus, African index Medicus, Google Scholar. Selected websites journals were also searched such as African index Medicus, African Journals OnLine (AJOL), Pan African Medical Journal (PAM), Annales Africaines de Médecine (AAM), Revue de Médecine et de Santé publique (RMSP) and Kisangani Medical .

The reference section of each of the articles included in the scoping review was also searched.

### Search

Words and phrases relevant to female breast and gynaecologic cancers were searched separately as text word in each database and grey literature sources: The same approach was performed with subject headings corresponding to the identified words and phrases. The words and phrases included: “female breast tumor” “female breast cancer”, “female breast neoplasm” “cervix cancer”, “cervical cancer “, “ovarian neoplasm”, “ovarian tumor “, vaginal cancer”, “tumor of the vagina”, “vulva tumor”, “vulvar neoplasm”, “vulvar cancer”, “uterine cancer”, “ endometrial neoplasm”, “uterus neoplasm”, “ Fallopian Tube Neoplasm”, “Democratic Republic of the Congo”, “ Belgian Congo” “Zaire”.

The retrieved articles were screened against the eligibility criteria and the purpose of the scoping review. This process was performed in two steps starting with title and abstract screening and repeated with the full text of the selected articles.

### Selection of sources of evidence

The initial selection was done by one reviewer (AK). The other co-authors reviewed the selection at each step and provided their approval to the next steps till the final selection of relevant articles. To be included in the review, articles needed to focus on one or more areas of cancer continuum regarding female breast cancer and/or gynecologic cancers, be published between January 2001 and April 2021, be written in English or French and report on studies that were either conducted in DRC, used data/samples from DRC.

### Data charting

A data charting form was used to extract relevant information from the identified literature. The following data were considered: authors’ names, year of publication, city and/or province where the study was conducted, study title, study design, areas of cancer continuum of care covered by the study objectives, key findings/results and language in which the article was written. The initial data charting was conducted by one author (AK). Data charting was then submitted to discussion and approval by all co-authors .

In this scoping review, the areas of cancer continuum include risk assessment, primary prevention, detection (screening), diagnosis (imaging, biopsy, laboratory tests),treatment, survivorship, and end-of-life care (15).

The areas of cancer continuum were defined as follows:

- Risk assessment : Evaluation of a person’s risk for cancer based on factors such as age, sex, personal and family medical history, ethnic background, lifestyle, genetics, exposure and screening history.
- Primary prevention: measures aimed to prevent cancer through the promotion of health and reduction of risks that are related to cancer occurrence.
- Detection: includes early diagnosis (or downstaging) and screening. Early diagnosis focuses on detecting symptomatic patients as early as possible, while screening consists of testing healthy individuals to identify those having cancers before any symptoms appear.(16)
- Diagnosis: processes that allow the identification of cancer from its signs and symptoms including laboratory tests, imaging, biopsies or any other appropriate procedure.
- Treatment: includes surgery, chemotherapy, radiation therapy,targeted therapy, immunotherapy, stem cell or bone marrow transplant and hormone therapy.
- Survivorship: includes the physical, mental, emotional, social, and financial effects of cancer that begin at diagnosis and continue through treatment and beyond. The survivorship experience also includes issues related to follow-up care, late effects of treatment, cancer recurrence, second cancers, and quality of life(17)
- End-of-life care: Care given to people who are near the end of life and have stopped treatment to cure or control cancer. End-of-life care includes physical, emotional, social, and spiritual support for patients and their families. End-of-life care may include palliative care, supportive care, and hospice care.(17)

### Synthesis of results

The results of the retrieved articles were summarized narratively and organized according to cancer sites and areas of cancer control covered.

## Results

### Selection of sources of evidence

A total of 448 articles were retrieved including 123 duplicates (figure 1). Of the remaining 225 articles, 172 were rejected after screening titles and abstracts leaving 53 articles eligible for examination of full texts. Twenty-one articles were removed, and the 32 remaining ones were included in the review. Seven articles were retrieved from search of the reference section of selected articles and one article was suggested by one of the co-authors (see figure 1).

**Figure 1.**
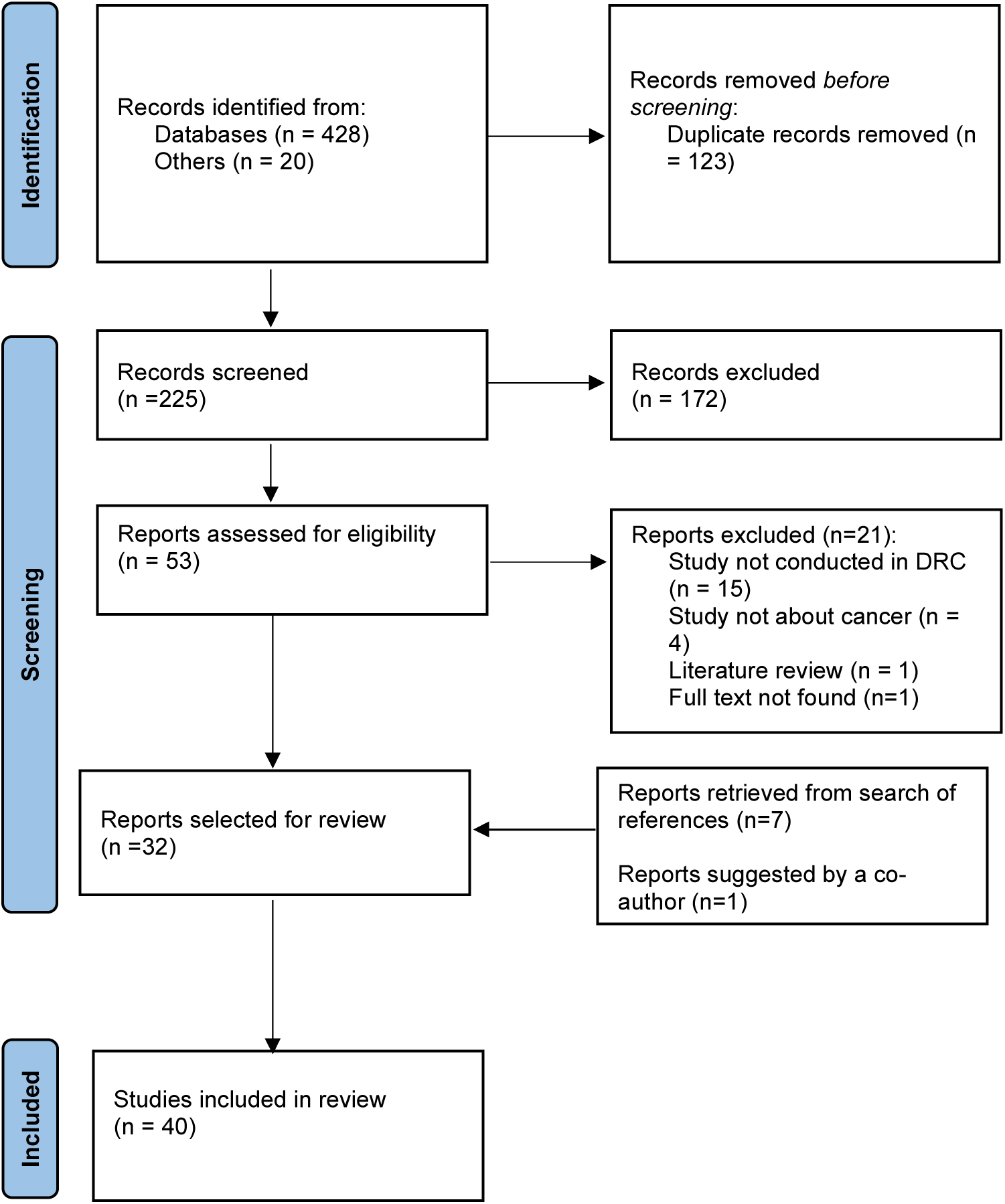
Selection of sources of evidence.

### Characteristics of sources of evidence

Twenty-one articles pertained to cervical cancer, 12 to female breast cancer and 2 to vulva cancer. Five articles focused on several cancer types. There was no article retrieved for the other gynaecologic cancers (uterine, Fallopian tube and ovarian cancers). Most of the studies (60%) used a cross-sectional design (see table 1).

**Table 1.** Study designs used in the selected sources of evidence.

| Types | N=40 | % |
| --- | --- | --- |
| Cross sectional | 24 | 60% |
| Qualitative studies | 2 | 5% |
| Randomized control trial (RCT) | 2 | 5 % |
| Case study | 4 | 10% |
| Case-control study | 1 | 2% |
| Cohort study | 1 | 2% |
| Cost-effectiveness studies | 2 | 5% |
| Retrospective study | 3 | 8% |
| Historical retrospective | 1 | 2% |

The sources of evidence revealed studies implemented in six DRC provinces. Most of the studies were conducted in Kinshasa (n=29). Two studies merged or compared data from two cities (Kinshasa and Lubumbashi). One study covered several sub-Saharan African countries including DRC. Figure 2 shows the different locations where studies were conducted.

**Figure 2:**
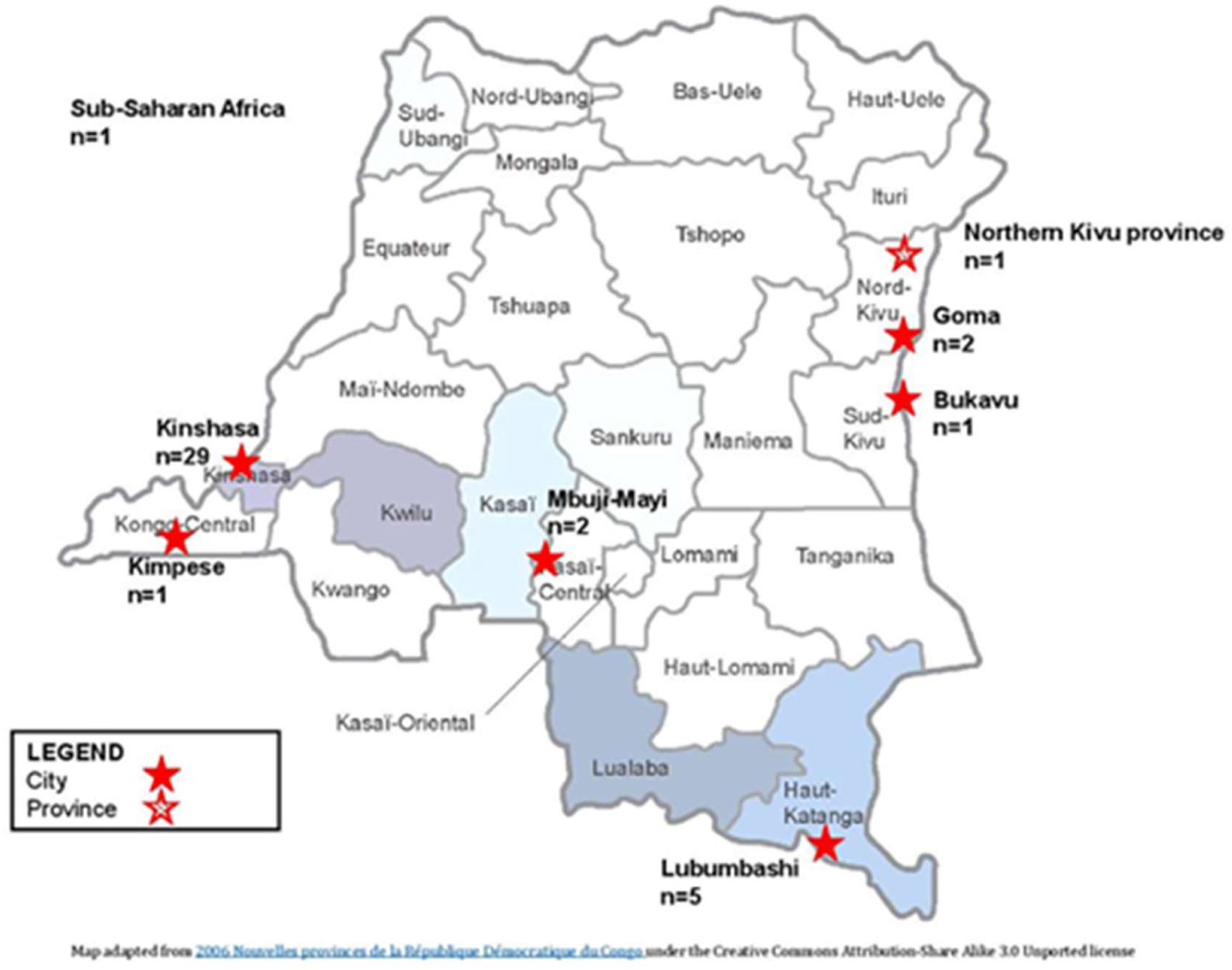
Locations of the selected studies.

### Cervical cancer

Out of the twenty-one articles focusing on cervical cancer, (see table 2).

**Table 2.**
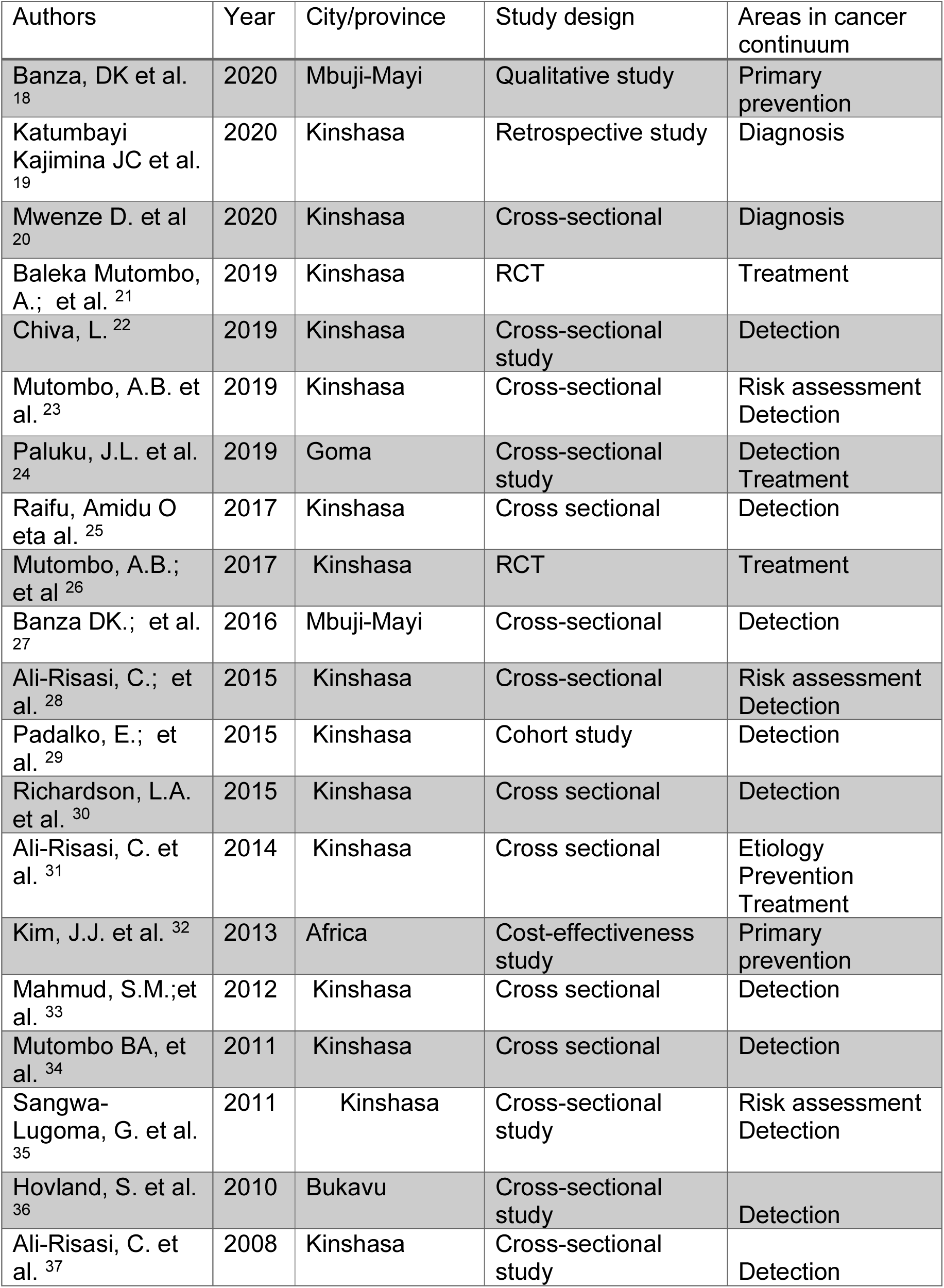
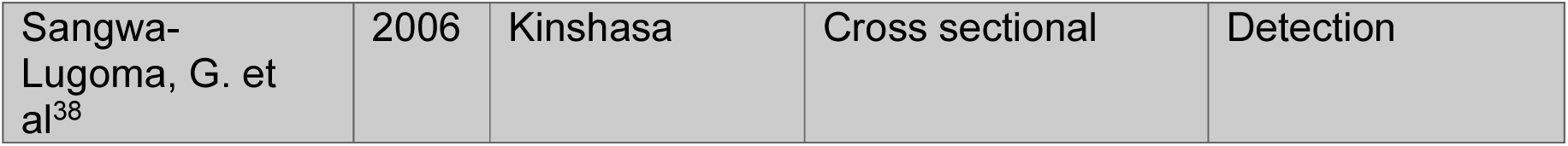
Studies about cervical cancer.

#### Risk assessment

Three studies focused on risk assessment in relation with cervical cancer (23,28,35). Regarding age, Mutombo, A.B. et al. (23) reported that human papilloma virus (HPV) prevalence was higher in women younger than 30 years old as well as in women aged 60 years and above. On the other hand, Sangwa-Lugoma G. et al (35) found that High risk -HPV (HR-HPV) prevalence was higher in women aged less than 35 years and decreased with age (35). One study (28) revealed a non statistically significant relationship between the number of sexual partners (≥3) and the prevalence of Low-grade Squamous Intraepithelial Lesions (LSIL). However, another study (35) showed that women whose partners had sex with prostitutes, used medical contraceptives smoke or were in a polygamous marriage had a statistically higher prevalence of HR-HPV. Women with higher socio-economic status had lower prevalence of HR-HPV (35). Women whose HIV status was unknown and who were using plants for vaginal care had higher prevalence of LSIL. HIV-positive women had a higher prevalence of LSIL than HIV-negative women did (28).

#### Prevention

Three articles looked at primary prevention of cervical cancer (18, 31, 32). One of them revealed that women and nurses had a limited knowledge of cervical cancer .Practitioners in Mbuji-Mayi had adequate theoretical knowledge of the disease but few opportunities related to practice (18).

A similar study conducted in Kinshasa reported that 82% of the participants had heard about cervical cancer (31) and 58% knew about early detection of cervical cancer lesions (31). A cost effectiveness analysis showed that 10.13 cases could be averted per 1000 vaccinated in the DRC (32).

#### Detection

Fourteen articles reported on detection of cervical cancer (22, 23, 24, 27-32, 33, 34, 35-38).

Three articles focused on visual inspection methods of cervical cancer detection with either acetic acid (VIA) or Lugol’s iodine (VILI). Paluku et al. (24) Reported that 7.5% of the women screened using VIA or VILI had lesions suggestive of cervical cancer. In Banza et al. (27) study, 38% of the VILI and VIA examinations performed were positive while Mutombo et al (34) obtained 20% using VIA. This latter study also revealed that VIA demonstrated high sensitivity, positive and negative predictive values compared to colposcopy. Six studies were based on HHV detection. Chiva et al (22) reported an HPV prevalence of 18% and they did not find HPV-18 in their sample. The prevalence of high-risk HPV was 23% in Mutombo et al (23) study and 12.5% in Sangwa-Lugoma et al (35). Padalko et al. (29) focused on searching HPV-53 prevalence and this latter was found in less than 1% of the samples (1/660).

Ali-Risasi et al. (37) found that the most prevalent HPV types were: 68, 35, 51, 52, 16, 31, 18, 17, 33, 45, 56, 58 and 59. HPV types 16 and 18 were found in 33% of the samples.

Hovland et al. (36) found that HPV detection methods were more sensitive that cytological techniques for cervical intraepithelial neoplasia grade 2 or worse.

Richardson et al (30) study revealed that the accuracy of Pap cytology decreased when the technicians were aware of patient’s HPV status. These technicians’ accuracy in Pap cytology in was compared between original readings and rereads.

Ali-Risasi et al. (28) used both Pap test and HPV detection. They found that the prevalence of LSIL+ was 31% in HIV positive women and 4% in HIVnegative women. The most frequent HPV types in HIV positive women were HPV 68, 35, 52,35, 51, 16. HPV 18 was observed in 1%. In HPV negative women or unknown HPV status, the most prevalent were in descending order HPV52, HPV35, HPV18, HPV51, HPV54, and HPV16 (n = 3), Mahmud et al. (33) reported in their study involving unscreened women in Kinshasa, DRC that HPV DNA assays (HC2, HC 2+4) demonstrated higher sensitivity and lower specificity compared to Pap test with regard to detecting cervical neoplasia .

Sangwa-Lugoma et al. (38) found similar results when comparing VIA and VILI performed by nurses and physicians to Pap cytology.

#### Diagnosis

Katumbayi Kajimina JC et al. (19) reported that immunostaining was significantly associated with the grade of neoplasia for p16 and for Ki-67 Mwenze D. et al (20) reported the lack of uniformity of PD-L1 expression in cyto-histological alterations predictive of HPV infection in neoplasia while it was highly expressed in high-grade intraepithelial neoplasia.

#### Treatment

Baleka Mutombo et al. (21) reported that the difference between the topical application of AV2 to cervical precancerous lesions was not significant compared to placebo. Another study (26) was about the registration of a randomized controlled trial exploring the efficacy of AV2 in the treatment of precancerous cervical lesions.

### Female breast cancer

Twelve articles reported on female breast cancer,

| Authors | Year | City/province | Study design | Areas in cancer continuum |
| --- | --- | --- | --- | --- |
| Henry-Tillman, R. et al <sup>39</sup> | 2021 | Kinshasa | Cross-sectional study | Detection |
| Kabongo MM et al. <sup>40</sup> | 2021 | Kinshasa | Cross-sectional study | Detection<br>Treatment |
| Parham, G.P. et al. <sup>41</sup> | 2021 | Kinshasa | Cross-sectional study | Detection<br>Diagnosis<br>Treatment |
| Irabor, O.C.; et al. <sup>42</sup> | 2020 | Kinshasa | Cost-effectiveness study | Treatment |
| Malingisi, G. Et al. <sup>43</sup> | 2020 | Kinshasa | Retrospective study | Risk assessment<br>Diagnosis |
| Mbala, K. ; et al <sup>44</sup> | 2019 | Kinshasa | Cross-sectional study | Risk assessment<br>Diagnosis |
| Nday Guy, et al <sup>45</sup> | 2018 | Kinshasa<br>Lubumbashi | Cross-sectional study | Diagnosis |
| Luyeye Mvila, G. et al. <sup>46</sup> | 2015 | Kinshasa | Case-control study | Diagnosis |
| Luyeye Mvila, G. et al. <sup>47</sup> | 2014 | Kinshasa | Cross-sectional study | Diagnosis |
| Nday G. et Al <sup>48</sup> | 2014 | Kinshasa<br>Lubumbashi | Cross-sectional study | Detection<br>Diagnosis |
| Ngombe, LK et al<br>49 | 2014 | Lubumbashi | Case study | Diagnosis |
| Lingohr, P. et al<br>50 | 2009 | Province Northern Kivu | Case study | Diagnosis |

#### Risk assessment

Two articles focused on female breast cancer risk assessment in Kinshasa (43,44).

Malingisi et al. (43) reported that women’s mean age at female breast cancer diagnosis was 47.50±10.75 years while Mbala et al (44) had 48,5 ± 10,2 years.

Both studies also found that most of the patients with female breast cancer were married, multiparous and in the premenopausal stage of their life (43–44).

#### Detection

Seven articles covered different aspects of female breast cancer detection. Henry-Tillman, R.et al (39) ; Kabongo MM et al.(40) and Parham, G.P. et al. (41) reported interventions conducted in a Kinshasa to improve health professionals capacity in female breast cancer detection . The training covered topics such as clinical female breast examination and female breast ultrasound diagnostics, ultrasound-guided core needle biopsy/fine needle aspiration (FNA) and female breast surgery. This training resulted in increased competency. Nday G.et Al (48) reported that female breast cancer cases were mostly diagnosed at stages T3 and T4.

#### Diagnosis

Parham, G.P. et al. (41) reported that local health professionals were trained in performing ultrasound-guided core-needle biopsy. Malingisi, G.et al.(43) and Mbala, K et al (44) found that the majority of female breast cancer were invasive ductal carcinoma. Nday Guy, et al (45) reported a statistically significant relationship between the tumor stage and Ki-67 value. Ki-67 positive value was more frequent for T3 and T4 tumors as well as G3. Luyeye Mvila, G. et al. (46) found that female breast cancer in a sample of women living in Kinshasa occurred in younger women and was mostly estrogen receptor negative and human epidermal growth receptor 2 positive. In another article, Luyeye Mvila, G. et al. (47) revealed that most of the female breast cancer were diagnosed at stage III and were invasive female breast cancer. They also identified BRCA gene mutation with history of female breast cancer occurrence in women of younger age.

Nday G.et Al (48) found that female breast cancer that were diagnosed in Kinshasa and Lubumbashi University hospitals were mostly invasive ductular carcinoma and grade 3 tumors. Tumor necrosis was observed in approximately one third of the biopsies. In addition, two studies reported cases of neoplastic pleurisy secondary to female breast cancer associated with lymphedema (49) and bilateral gigantic Burkitt’s lymphoma of the female breast (50)

#### Treatment

Two studies (40–1) reported that local health professionals were trained in female breast surgery and administration pf chemotherapy for female breast cancer. These professionals performed different surgical procedures such as lumpectomies, radical mastectomies, axillary lymph node dissections, oncoplastic surgery and palliative resections. Only few minor complications wee observed (wound dehiscence). Irabor, O.C.; et al. (42) found that the projected national cost savings with hypofractionated female breast radiation therapy in DRC (2019–2025 be US$ 40.4 million and the projected national cost for full access to female breast radiation therapy at conventional fractionation in DRC (2009–2025) US$ 180 million.

#### Vulva cancer

Two articles focused on vulva cancer. Both pertained to the diagnosis of this cancer through biopsy followed by treatment (vulvectomy and lymphadenectomy). Mukeya, G.K. et al (51) reported a case in a woman in a 72-year old woman while Idy Y.I et al. (52) described a case in 32-year old woman .

#### Multiple sites

Female breast cancer and cervical cancer Risk assessment /Diagnosis

Mashinda DK et al. (53) conducted a qualitative study to explore cancer patients’ journey. They found that this latter was long, expansive.

Metaphysics was frequently related to the occurrence of cancers.

#### Treatment

Gombo Y et al (54) reported their experience in establishing a chemotherapy infusion unit and training Congolese healthcare providers in chemotherapy treatment of cervical and female breast cancers. While Hicks, M.L. et al. (55) present their approach in building capacity of local healthcare providers in surgical treatment of cervical cancer.

#### Female breast/cervical/uterine Diagnosis

Lukanu N.P et al (56) in their epidemiological study of cancer in in a rural hospital in DRC found that among female patients, a predominance of female breast cancer was the most prevalent (41.3%), followed by cervical cancer (21.4%), endometrial cancer (6 %), ovarian (3.2%), vulva (1.1 %) and vagina (0.7%).

Kalisya, L.M et al. (57) through a study conducted in a hospital located in the Eastern region of DRC reported that invasive female breast ductal carcinoma prevalence represented 2.9% of observed female breast lesions. In addition, cervical and uterine lesions were observed in approximately 40% of the sample.

## Discussion

This scoping review revealed several gaps in research focusing on female breast cancer and gynaecological cancers in the Democratic Republic of the Congo. Studies were predominantly reporting on cervical and female breast cancers. They were conducted in a limited number of regions and cities of the country; they covered only few areas of the cancer continuum of care and used a limited number of study designs.

More precisely, most studies were conducted in Kinshasa, and only six provinces out of the existing 24 were involved in the last 20 years covered by the literature search. The review demonstrated that most of the studies were cross-sectional. There were no studies about survivorship and end-of-life care.

These findings are consistent with studies conducted in other African countries. For instance, a literature review reported that cancer research conducted in Zambia has similarly focused mainly on cervical cancer (58). Another review showed that in Nigeria, most breast cancer studies were cross-sectional and case report studies (59). A review of cancer research in Kenya revealed that studies were conducted in limited regions and types of setting. In addition, cervical cancer represented approximately 35% of cancer research followed by breast cancer (12%) whereas one percent for gynecologic cancers represented only 1% of cancer research. Finally, studies used predominantly observational study designs and focused mainly on early detection, diagnosis, and prognosis. (60)

Regarding cervical cancer, many studies described detection -related findings using a variety of techniques. The only article tackling treatment assessed an experimental treatment that was not conclusive. Only few articles studied risk assessment, prevention, and diagnosis. As for female breast cancer, the literature weighted towards detection and diagnosis. There was a limited number of articles about risk assessment and few focused on treatment. In twenty years, only two articles pertaining to vulva cancer were retrieved. Only one study provided a quite thorough epidemiologic overview of female breast and gynaecological cancers in a rural hospital.

This scoping review shows that there is a clear need to not only broaden the scope of female breast and gynaecologic cancer research but also to build the capacity in research focusing on these conditions.

Specifically, training in not only female breast and gynaecological cancers but also in cancer research methodologies should be provided to practising and in-training health professionals in DRC.

Cancer risk assessment- and primary prevention-related research and clinical skills need to be developed to improve the outcomes of these cancers. Risk assessment and primary prevention in a limited-resources country such as DRC is particularly needed to avoid the economic and social costs of advanced cancers that are not at all bearable by the society and women.

Detection and diagnosis although extensively covered by articles retrieved through this review need to be taught and implemented according to up-to-date guidelines such as for instance the WHO guideline for screening and treatment of cervical pre-cancer lesions for cervical cancer prevention (53) .

Efficient cervical cancer prevention through HPV immunization has been proven and is implemented in few countries in Africa. Advocacy and research in this field are needed to move the agenda of HPV immunization in DRC.

Cancer treatment is costly and not affordable or even existing in several parts of the country. Again research, training and advocacy are needed in this area.

The lack of studies on cancer survivorship and end-of-life care in DRC is not surprising as most of the cases are identified at an advanced stages and treatment is often limited to surgery that have limited impact on extending patients survival and quality of life. Consequently, there is a need for more research not only to better understand the outcomes and experiences of these patients but also to develop the delivery of quality palliative care. Palliative care can be delivered at home or in hospital settings and this is applicable in resource-limited environment as well.

An adapted and realistic approach needs to be developed to address the gaps and needs identified through this review. Five strategies need to be prioritized.

1. Female breast and gynaecological cancers research priorities need to be identified.
2. A realistic research agenda needs to be established based on identified needs and priorities
3. Relevant stakeholders from different sectors need to be involved including providers, researchers, policy makers, patients, families, and organizations representing them.
4. An adapted research and education training program needs to be developed
5. An advocacy plan needs to be developed.
6. Strategies to improve funding for research both at the national and international level need to be elaborated.

## Limitations

The main limitation of this study is due to the fact that we conducted only an online search of literature. Although grey literature was considered, we did not have access to paper-based research conducted in Congolese or other institutions. Future research should consider this issue in their literature search strategy.

## Conclusion

This scoping review has demonstrated significant gaps in female breast and gynaecologic cancer research in DRC. The review’s findings support the need for further research in all areas of the continuum of cancer care, the establishment of a clear and adapted research agenda, advocacy and providers’ capacity development. Consequently, the DRC National Center for the Fight against Cancer is currently planning as immediate next step to clarify research priorities in female breast and gynaecologic cancers in collaboration with relevant stakeholders.

## Data Availability

All data produced in the present work are contained in the manuscript.

